# Identification of Blood-based Non-invasive Biomarkers and Therapeutic Agents against Pancreatic Ductal Adenocarcinoma (PDAC): A Network-based Study

**DOI:** 10.1101/2020.01.08.20016931

**Authors:** Md. Asad Ullah, Bishajit Sarkar, Fayza Akter

## Abstract

Pancreatic Ductal Adenocarcinoma (PDAC) is the most demolishing form of pancreatic cancer with poor prognosis and rising incidence. Difficulties in the early detection and aggressive biological nature of this disease are responsible for most of the therapeutic failures. In this study publicly available microarray expression data of full RNA from peripheral blood of PDAC patient has been utilized via network-based approach in order to identify potential non-invasive biomarkers and drug targets for early diagnosis and treatment of PDAC. Analysis of differentially expressed genes revealed their predominant involvement in translational process, apoptotic process, protein phosphorylation, immune responses, ATP binding, protein binding and signal transduction. Moreover, CREBBP, MAPK14, MAPK1, SMAD3, UBC, MAGOH, HSP90AB1, RPL23A, ACTB and STAT3 were identified as the best proteome signatures, GATA2, FOXC1, PPARG, E2F1, HINFP, USF2, MEF2A, FOXL1, YY1 and NFIC were identified as the best transcriptional regulatory signatures, and hsa-miR-93, hsa-miR-16, hsa-miR-195, hsa-miR-424, hsa-miR-506, hsa-miR-124, hsa-miR-590-3p, hsa-miR-1, hsa-miR-497 and hsa-miR-9 were identified as the best post-transcriptional regulatory signatures in PDAC patient. Analysis of drug-gene interaction revealed Anisomycin, Azactidine, Arsenic trioxide, Bortezomib, Ulixertinib and some other molecules as the probable candidate molecules which may reverse PDAC condition.

## 1. Introduction

Pancreatic Ductal Adenocarcinoma (PDAC) is by far the most common type of devastating exocrine neoplastic and chemo resistant cancer, concerning 90% of all pancreatic cancers (Orth et al., 2019). This cancer originates in the ducts that carry secretions of digestive enzymes and bicarbonate away from the pancreas. Pancreatic cancer is rarely diagnosed before 55 years of age, mainly the highest incidence is reported in people over 70 years (Siri and Salehiniya, 2019). Though, there has no evidence what causes this cancer in most of cases, many researchers are identified some genetic and environmental factors (obesity, smoking, diabetes etc.) are intertwined in the development of PDAC. This cancer occurs when cells in our pancreas especially in the duct line develop mutations in the DNA that causes cells to grow uncontrollably and to continue living further can form a tumor. Malignancy can spread to nearby organs and blood vessels if the cell remains untreated for the long time (Adamska et al., 2017). Furthermore, 25% of people survive one year and 5% live for five years for the lack of apparent and particular symptoms and reliable biomarkers for early diagnosis as well as aggressive metastatic spread leading to poor response to treatments (Maitra and Hruban, 2008). Globally, 458,918 new cases of pancreatic cancer have been reported in 2018, and 355,317 new cases are estimated to occur until 2040 (Rawla etal., 2019). By 2030, researchers project that pancreatic cancer will become the 2nd leading cause of cancer related death in the US after lung cancer, surpassing colorectal, breast, and prostate cancer (Rahib et al., 2014). Moreover, currently accessible therapeutic options are surgery, radiation, chemotherapy, immunotherapy, and use of targeted drugs. But these are not effective as later stage detection causes metastasis additionally it is expensive too (Charsi, 2007; Orth et al., 2019). Therefore, there is an increasing demand for the development of novel, effective strategies aiming to advance current therapeutic possibilities.

Microarray data is now being increasingly used to assume the function of differentially expressed genes (DEGs) which are not currently available (Ullah et al., 2019; Hatfield et al., 2003). And DEGs identified from microarray data can be used in the robust identification of biomarkers. However, these studies provide fruitful findings but the understanding of actual mechanism of biological condition using the data of differentially expressed genes (DEGs) is often difficult and may sometime come with erroneous interpretation (Pepe et al., 2003; Crow et al., 2019).

In this study we have employed network-based approach to identify potential molecular signatures which could be used as biomarkers or drug targets in the early diagnosis and treatment of PDAC. We have also employed strategy to identify drug-gene interactions in PDAC in order to select feasible therapeutic molecule for the treatment of PDAC (**Figure 1**).

**Figure 1:**
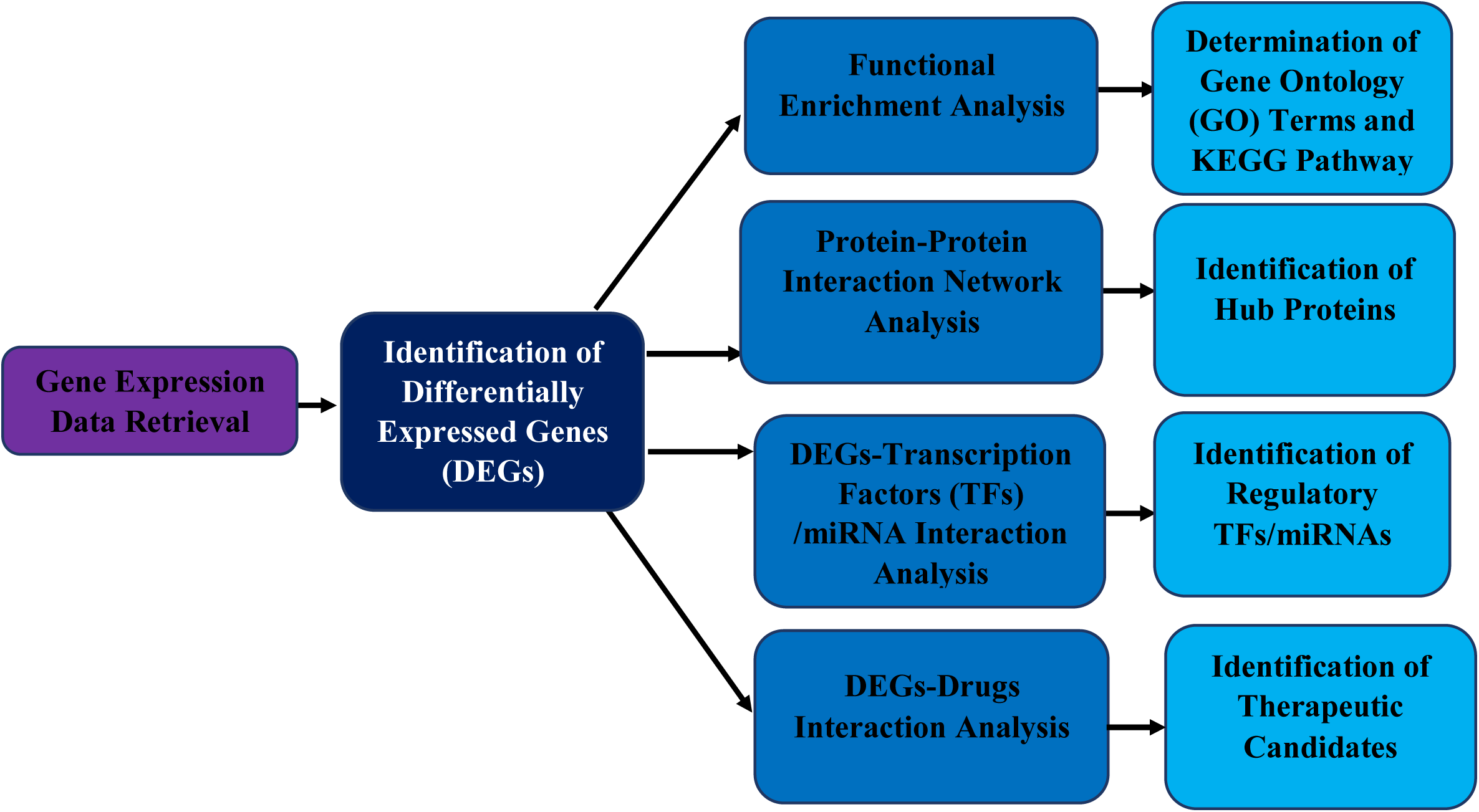
Strategies employed in the overall study.

## 2. Materials and Methods

### 2.1. Data Retrieval and Identification of Differentially Expressed Genes

We retrieved GSE74629 microarray data from NCBI-GEO (National Center for Biotechnology Information-Gene Expression Omnibus) database (Caba et al., 2015). The dataset comprises the expression profile of total RNA from peripheral blood of 36 PDAC patients and 14 age, gender and habit matched healthy people. The presence of PDAC was confirmed by histological biopsy or imaging-guided biopsy. After retrieval, the data was statistically analyzed using GEAP (Gene Expression Analysis Platform) to differentiate the upregulated and downregulated genes (Nunes et al., 2018). Log2 transformation was applied and differentially expressed genes (DEGs) were sorted with adjusted P value<0.01 filter since the lower value corresponds to more accurate prediction.

### 2.2. Functional Enrichment Analysis of DEGs

Both upregulated and downregulated gene sets were analyzed by DAVID (Database for Annotation, Visualization, and Integrated Discovery) (version 6.8) for gene over-representation to elucidate gene ontology (GO) terms and pathways involved with DEGs (Sherman and Lempicki, 2009). P values were adjusted using the Hochberg and Benjamini test and gene count >15 were set as the cut-off point during the analysis.

### 2.3. Construction of Protein-Protein Interaction Network and Identification of Hub Proteins

STRING (Search Tool for the Retrieval of Interacting Genes/Proteins) database was utilized for the construction of generic protein-protein interaction (PPI) network with NetworkAnalyst (Szklarczyk et al., 2017; Xia et al., 2015). Topological and expression analysis of the DEGs were performed using NetworkAnalyst. Hub proteins in the generic PPI network with top 10 most connected nodes were identified with cytoHubba plugin using betweenness centrality interaction matrix on Cytoscape (version 3.7.2) (Chin et al., 2014; Shannon et al., 2003). Then the functional enrichment of the hub proteins was also analyzed.

### 2.4. Identification of Regulatory Molecules

DEGs were searched against JASPAR which is an open access, curated and non-redundant database of DNA binding transcription factors, with the help of NetworkAnalyst to construct transcription factor (TF)-DEGs interaction network (Sandelin et al., 2004). Micro RNA (miRNA)-DEGs interaction network was constructed searching the DEGs against TarBase, a manually curated microRNA database that includes almost 1,300 experimentally supported targets (Sethupathy et al., 2006). Top 10 interacting TFs and miRNAs were selected and analyzed.

### 2.5. Identification of Small Therapeutic Candidate Molecules

After the identification of hub proteins and regulatory biomolecule signatures, the selected signatures were then analyzed by DGidb (Drug Gene Interaction Database) (version 3.0) database that presents drug-gene interaction and gene druggability information in order to identify potential therapeutic candidate molecules that may reverse the PDAC condition (Cotto et al., 2017). Best observed therapeutic molecules according to specific target were then selected.

## 3. Result

### 3.1. Transcriptome Signatures

Publicly available microarray data of total RNA profile from peripheral blood of PDAC patient and control was retrieved for statistical analysis. A total of 1910 differentially expressed genes were identified with 681 upregulated and 1229 downregulated genes were identified within the defined parameter (**Figure 2**).

**Figure 2:**
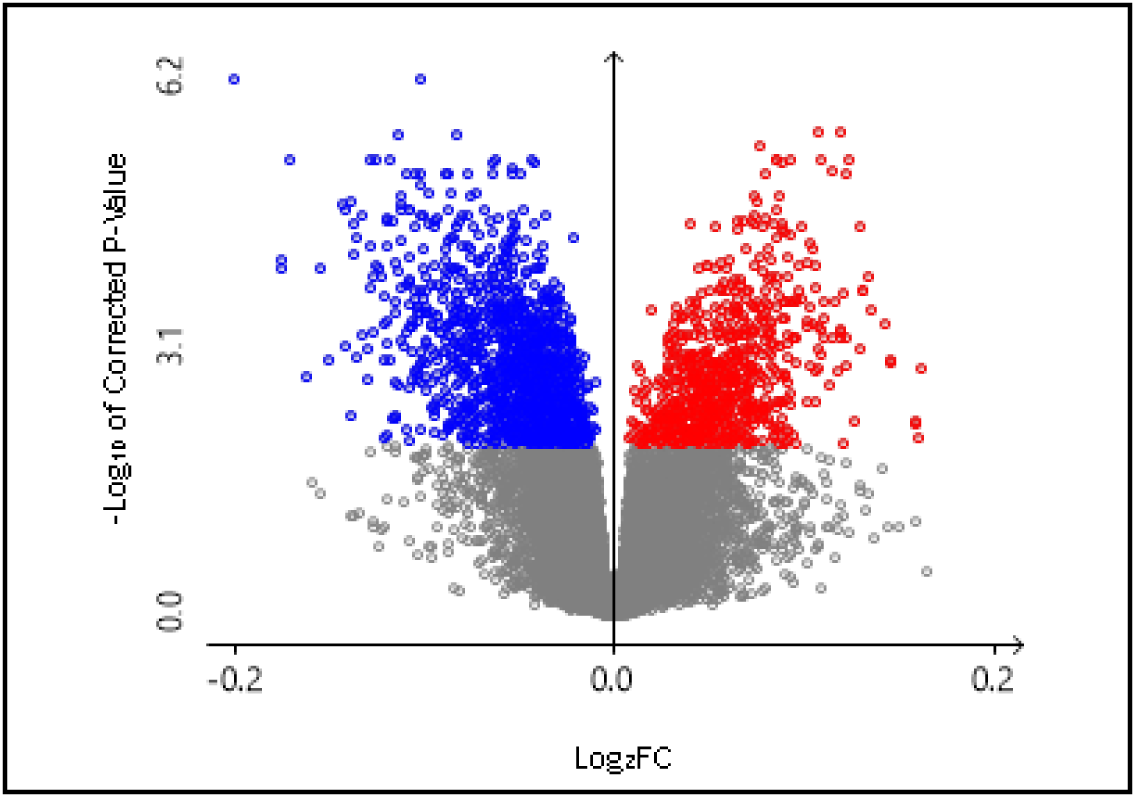
Volcano plot of selected differentially expressed genes (DEGs). Colored (Blue: Down regulated genes; Red: Upregulated genes) DEGs have been selected with adjusted P value > 0.01. filter.

After that both upregulated and downregulated gene sets were analyzed sequentially to understand their functional enrichment reflecting Gene Ontology (GO) terms and enriched pathway. GO aids in understanding the involvements of the genes in context of Biological Processes, Cellular Compartmentation and Molecular Function. Top 10 GO terms for both upregulated and downregulated gene sets were retrieved (**Table 1**). Among the identified terms of upregulated genes, 55 genes were found to be involved in signal transduction and 31 genes were involved in innate immune response. Again, Protein Phosphorylation, Apoptotic Processes and Viral Processes were the next predominant GO terms. Moreover, the upregulated genes were also predominant in ATP binding, protein binding and Protein homodimerization activity and they were shown to act mainly in cytoplasm, cytosol and plasma membrane.

**Table 1:**
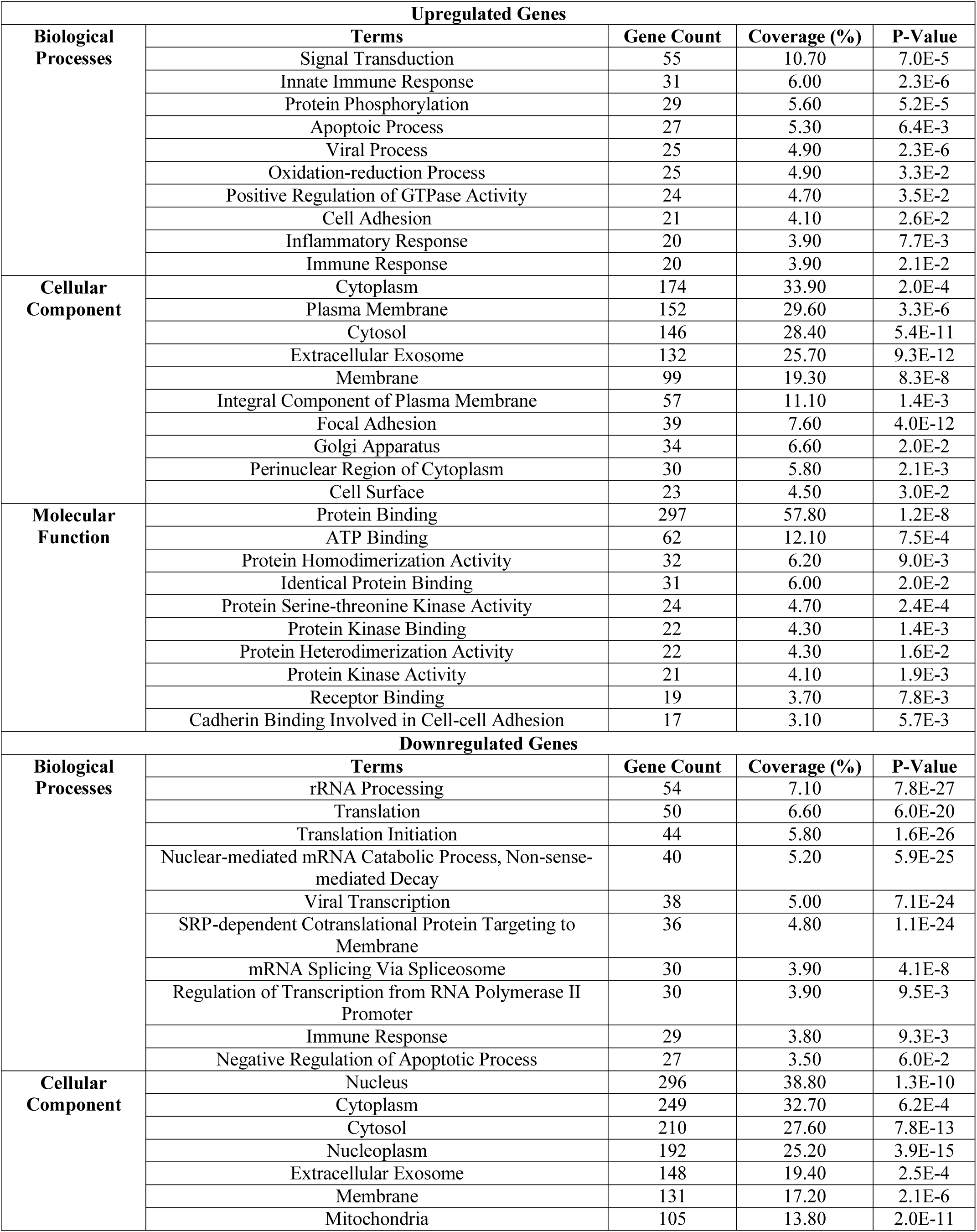
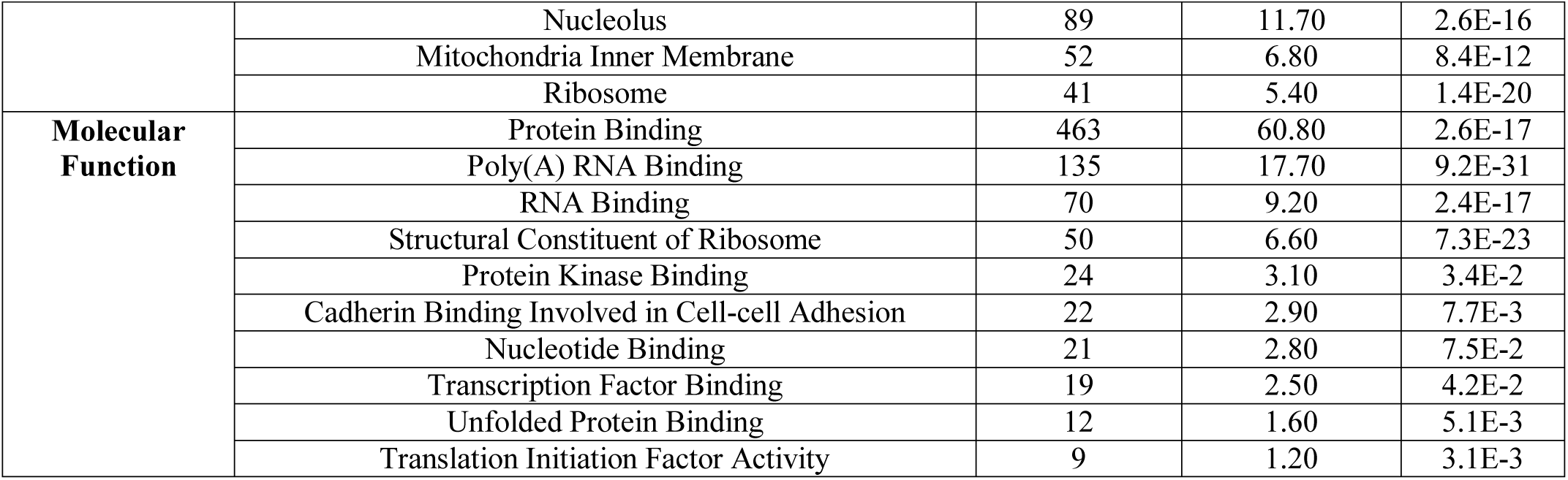
Top 10 gene ontology (GO) terms of differentially expressed genes (DEGs) in PDAC.

Among the downregulated genes, 54, 50 and 44 genes were found to be involved in rRNA processing, Translation and Translational Initiation respectively. Additionally, they were found to be involved in protein binding and RNA binding and also observed to act predominantly in cytosol and nucleus.

Moreover, The DEGs were then again subjected to analyze their involvement in biological pathway. Selected genes showed sign of their involvement in KEGG (Kyoto Encyclopedia of Genes and Genomes) pathway (**Figure 3**).

**Figure 3:**
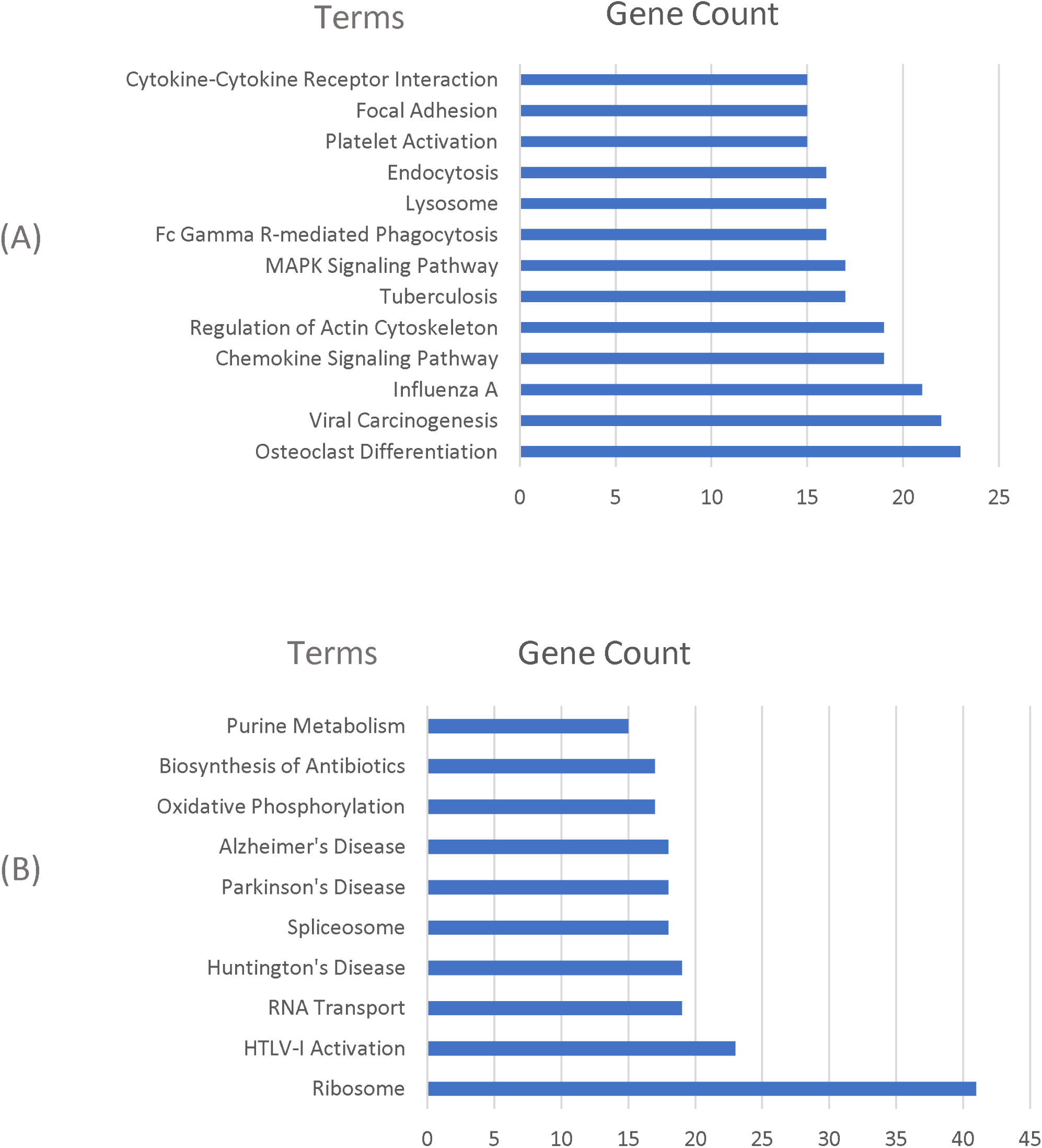
KEGG pathway of differentially expressed genes: (A) Upregulated genes; (B) Downregulated genes.

### 3.2. Proteome Signatures

After functional enrichment analysis, the DEGs were used to construct protein-protein interaction (PPI) network. A densely connected scale free network was constructed for the DEGs (**Figure 4**). Then the generic PPI map was used to identify the hub proteins using the betweenness centrality matrix. CREBBP, MAPK14, MAPK1, SMAD3, UBC, MAGOH, HSP90AB1, RPL23A, ACTB and STAT3 were identified as the most connected nodes (Hub proteins) (**Figure 5**) (**Table 2**). Then the functional enrichment of the hub proteins was analyzed. The identified proteins were primarily involved in positive regulation of protein transport into nucleus (P value: 8.30E-06), Regulation of immune response (P value: 1.32E-05) and posttranscriptional regulation of gene expression (P value: 1.32E-05). They were also found to be involved in phosphatase binding and performing predominantly in nucleus.

**Table 2:**
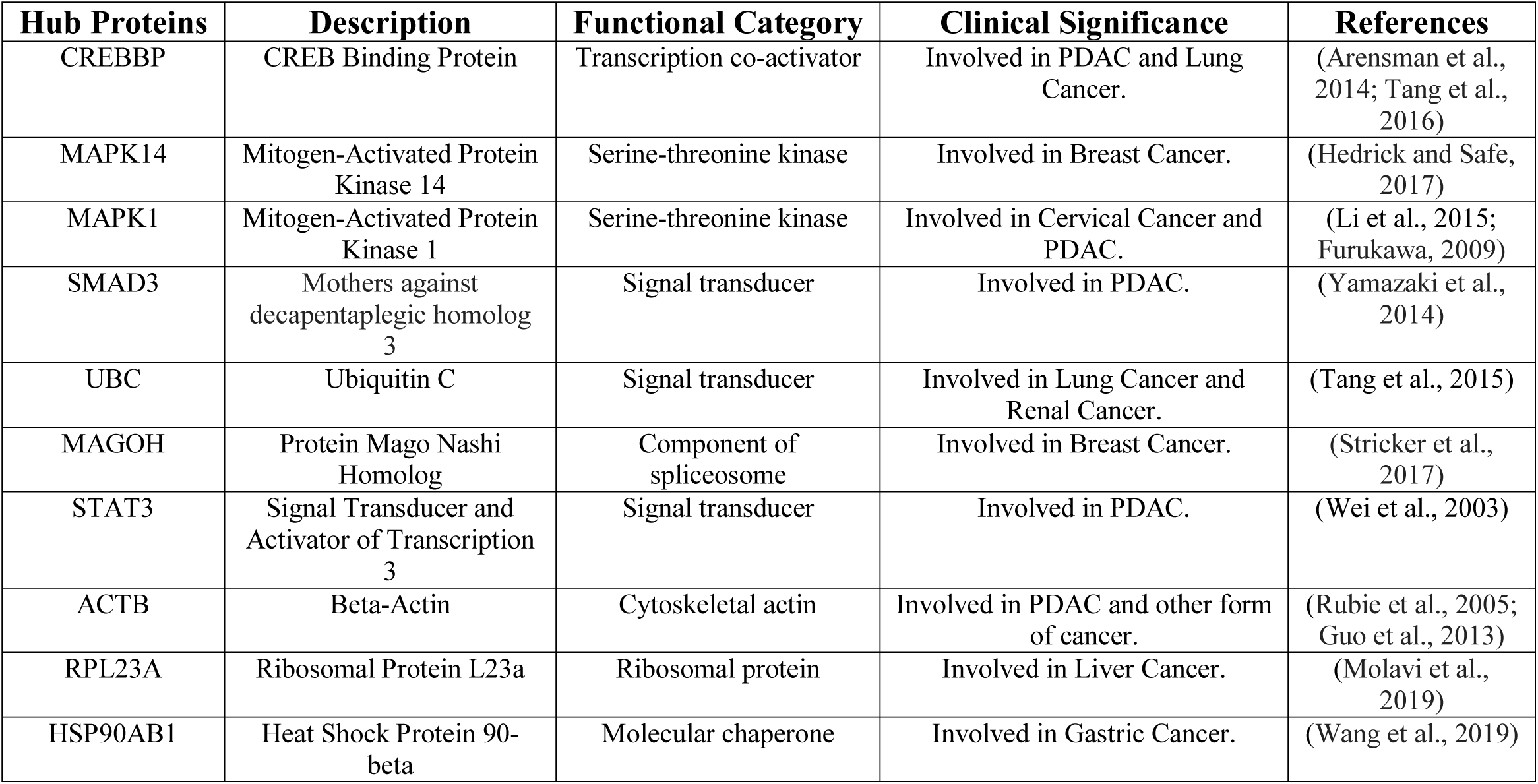
Summary of the identified hub proteins from PPI network.

**Figure 4:**
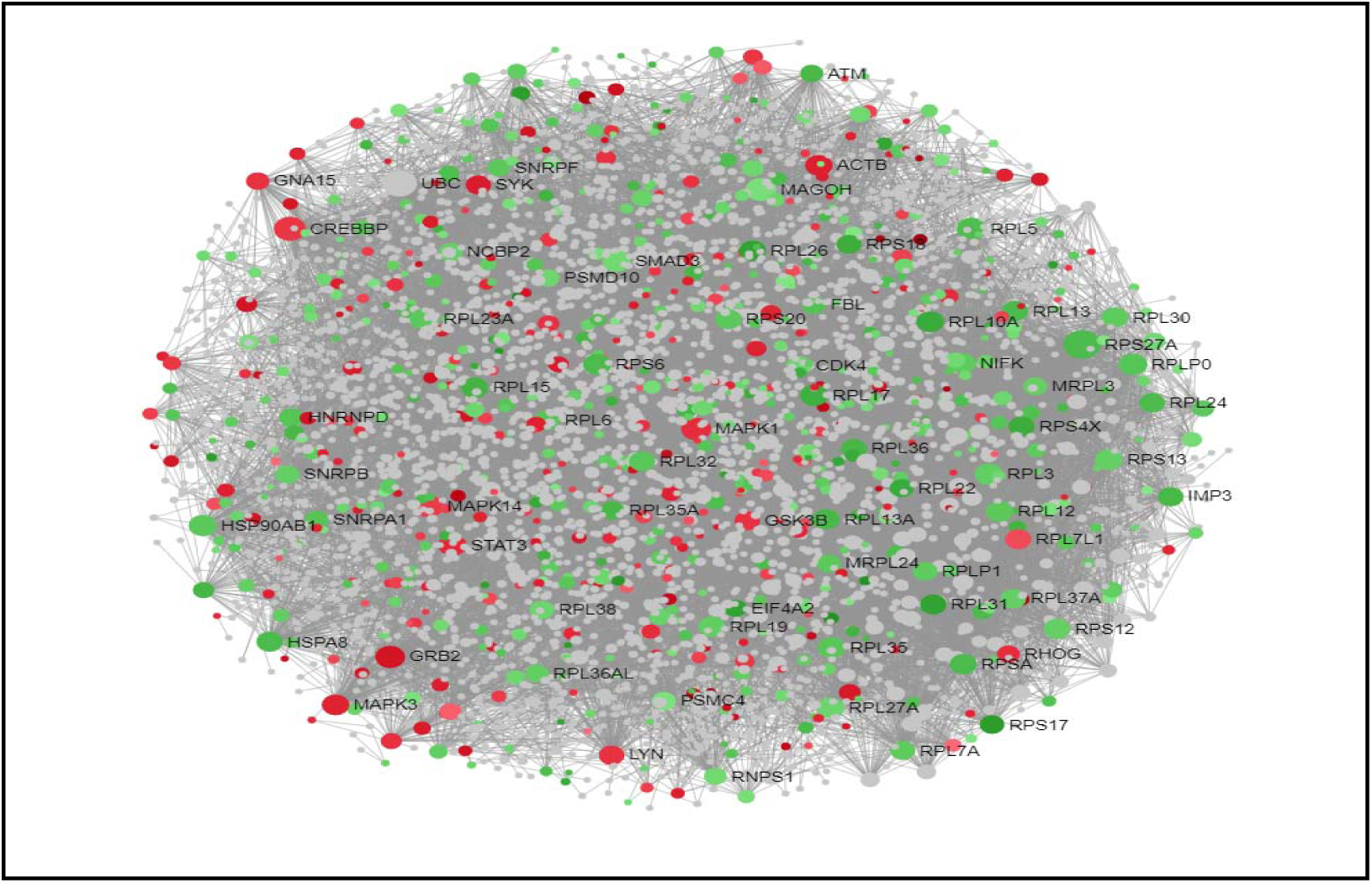
Protein-protein interaction (PPI) network of differentially expressed genes (DEGs). Nodes represent DEGs (Green: Upregulated genes; Red: Downregulated genes). Edges represent interaction.

**Figure 5:**
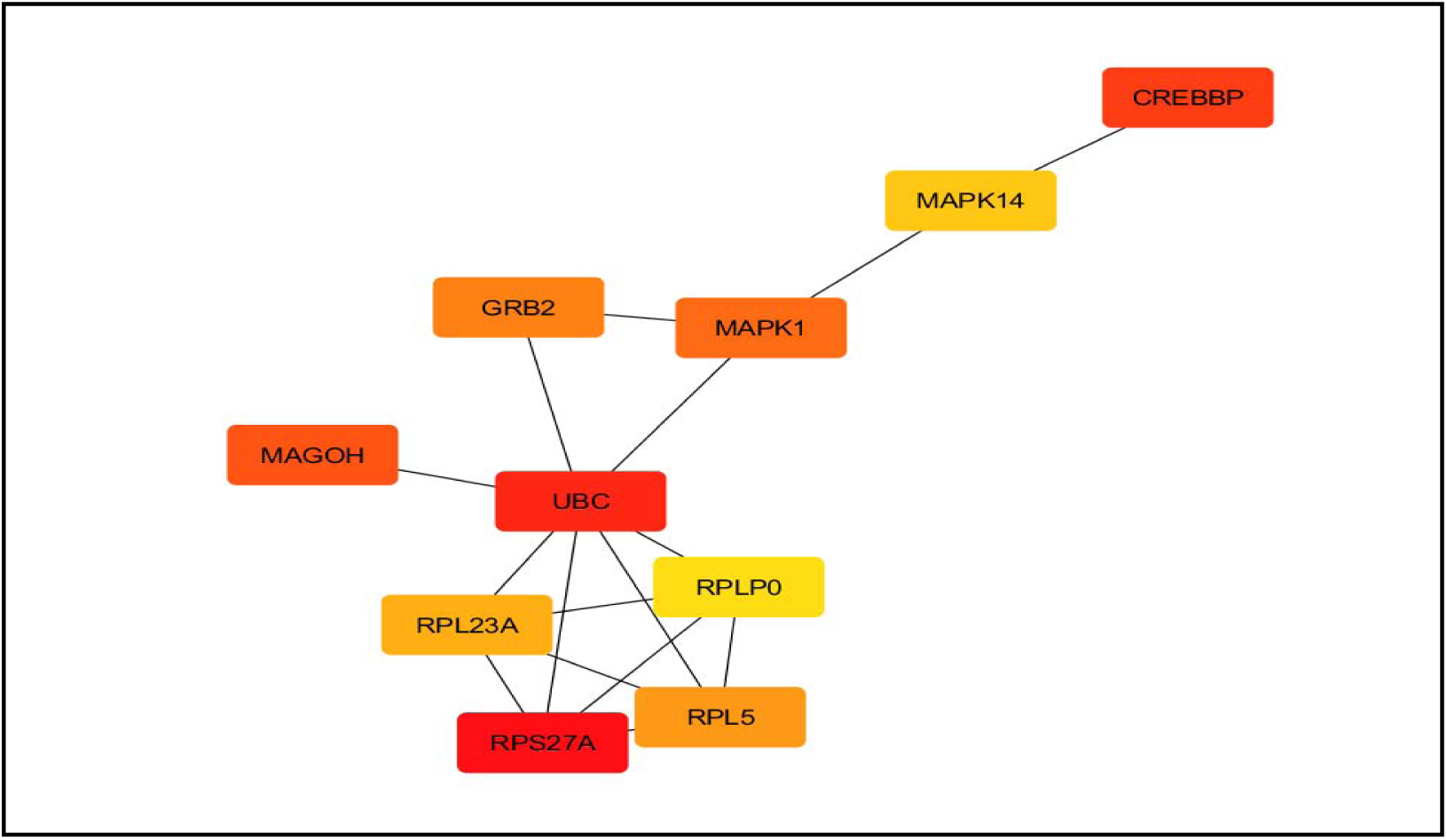
Hub proteins from generated protein-protein interaction (PPI) network. Nodes represent proteins and edges represent interactions.

### 3.3. Regulatory Signatures

The DEGs were analyzed to identify transcriptional (TFs) and post-transcriptional (miRNAs) regulatory biomolecules. GATA2, FOXC1, PPARG, E2F1, HINFP, USF2, MEF2A, FOXL1, YY1 and NFIC were identified as the best transcriptional (TFs) regulatory biomolecules (**Figure 6**) (**Table 3**). And, hsa-miR-93, hsa-miR-16, hsa-miR-195, hsa-miR-424, hsa-miR-506, hsa-miR-124, hsa-miR-590-3p, hsa-miR-1, hsa-miR-497 and hsa-miR-9 were identified as the best post-transcriptional (miRNAs) regulatory biomolecules (**Figure 7**) (**Table 4**).

**Table 3:** Summary of the selected TFs from TFs-DEGs network.

| **Transcription Factors** | **Description** | **Clinical Significance** | **References** |
| --- | --- | --- | --- |
| GATA2 | GATA Binding Factor 2 | Involved in Prostate Cancer. | (Chiang et al., 2014) |
| FOXC1 | Forkhead Box C1 | Involved in PDAC. | (Wang et al., 2013) |
| PPARG | Peroxisome Proliferator Activated Receptor Gamma | Involved in PDAC and other cancers. | (Wang et al., 2015; Elnemr et al., 2000) |
| E2F1 | E2F Transcription Factor 1 | Involved in PDAC. | (Schild et al., 2009) |
| HINFP | Histone Nuclear factor | Involved in different cancers. | (Holmes et al., 2005, van et al., 2009) |
| USF2 | Upstream Stimulatory Factor 2 | Involved in Lung Cancer. | (Ocejo-Garcia et al., 2005) |
| MEF2A | Myocyte enhancer factor-2A | Involved in Liver Cancer. | (Pon and Marra, 2016) |
| FOXL1 | Forkhead Box L1 | Involved in PDAC. | (Zhang et al., 2013) |
| YY1 | Yin Yang 1 | Involved in PDAC. | (Yuan et al., 2017) |
| NFIC | Nuclear Factor 1-C | Involved in Breast Cancer. | (Lee et al., 2015) |

**Table 4:** Summary of the selected miRNAs from miRNAs-DEGs network.

| **MiRNAs** | **Description** | **Clinical Significance** | **References** |
| --- | --- | --- | --- |
| hsa-miR-93 | *Homo sapiens* micro-ribonucleic acid -93 | Involved in Gastric Cancer | (Larki *et al.*, 2018) |
| hsa-miR-16 | *Homo sapiens* micro-ribonucleic acid -16 | Involved in PDAC | (Huang *et al.*, 2019) |
| hsa-miR-195 | *Homo sapiens* micro-ribonucleic acid -195 | Involved in Breast Cancer | (Singh *et al.*, 2015) |
| hsa-miR-424 | *Homo sapiens* micro-ribonucleic acid -424 | Involved in PDAC | (Lee *et al.*, 2007) |
| hsa-miR-506 | *Homo sapiens* micro-ribonucleic acid -506 | Involved in Cervical Cancer | (Li *et al.*, 2011) |
| hsa-miR-124 | *Homo sapiens* micro-ribonucleic-acid -124 | Involved in Nasopharyngeal Carcinoma | (Peng *et al.*, 2014) |
| hsa-miR-590-3p | *Homo sapiens* micro-ribonucleic acid -590-3p | Involved in Colorectal Cancer | (Elfar *et al.*, 2019) |
| hsa-miR-1 | *Homo sapiens* micro-ribonucleic acid -1 | Involved in PDAC and Colon Cancer | (Zhu and Wang, 2016; Xu *et al.*, 2015) |
| hsa-miR-497 | *Homo sapiens* micro-ribonucleic acid -497 | Involved in different cancer | (Wu *et al.*, 2016) |
| hsa-miR-9 | *Homo sapiens* micro-ribonucleic acid -9 | Involved in Lung Cancer | (Han *et al.*, 2017) |

**Figure 6:**
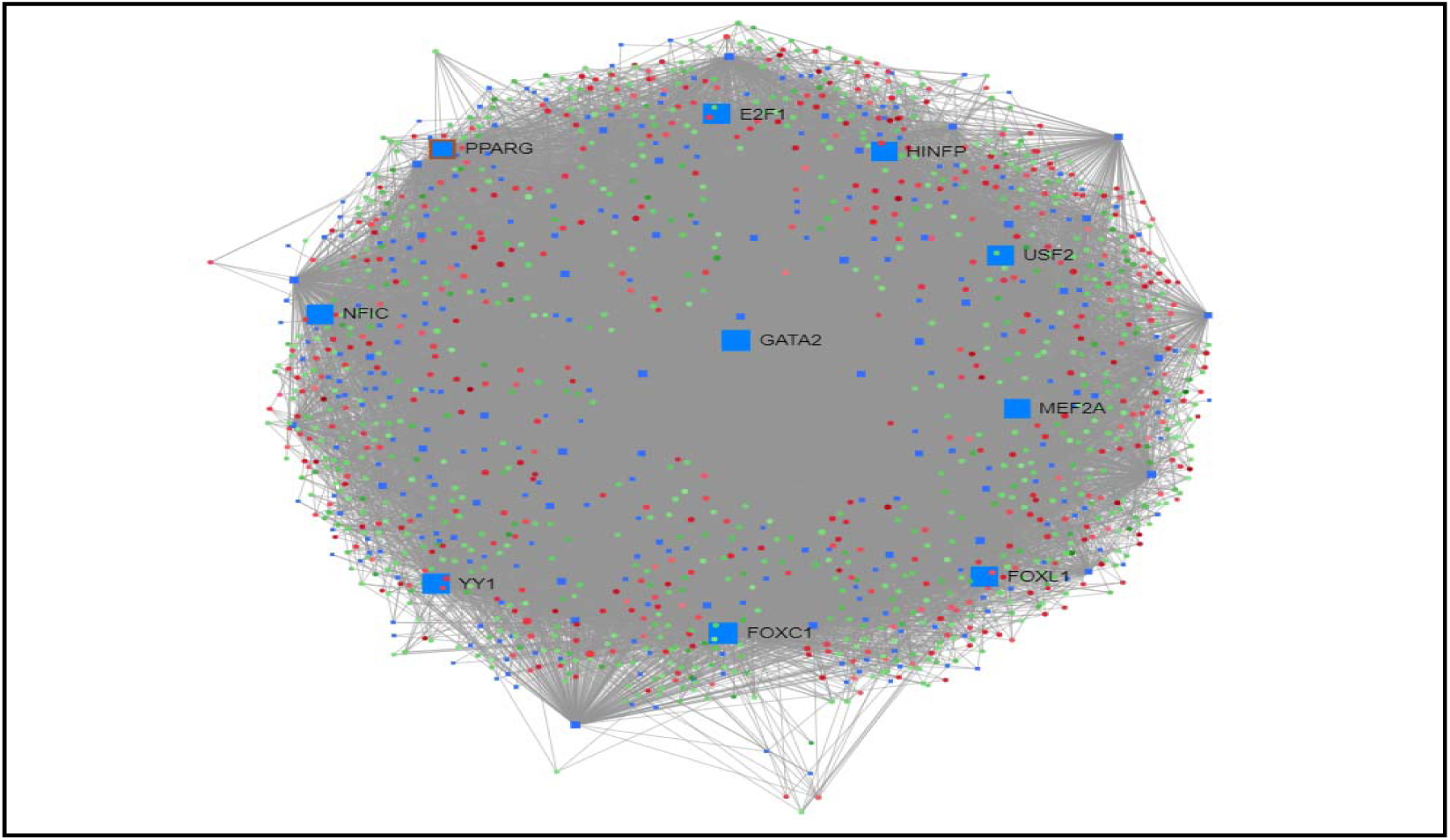
Interaction between transcription factor (TF) and differentially expressed genes network of differentially expressed genes (DEGs). Nodes represent DEGs (Green: Upregulated genes; Red: Downregulated genes; Blue: Transcription factors). Edges represent interaction.

**Figure 7:**
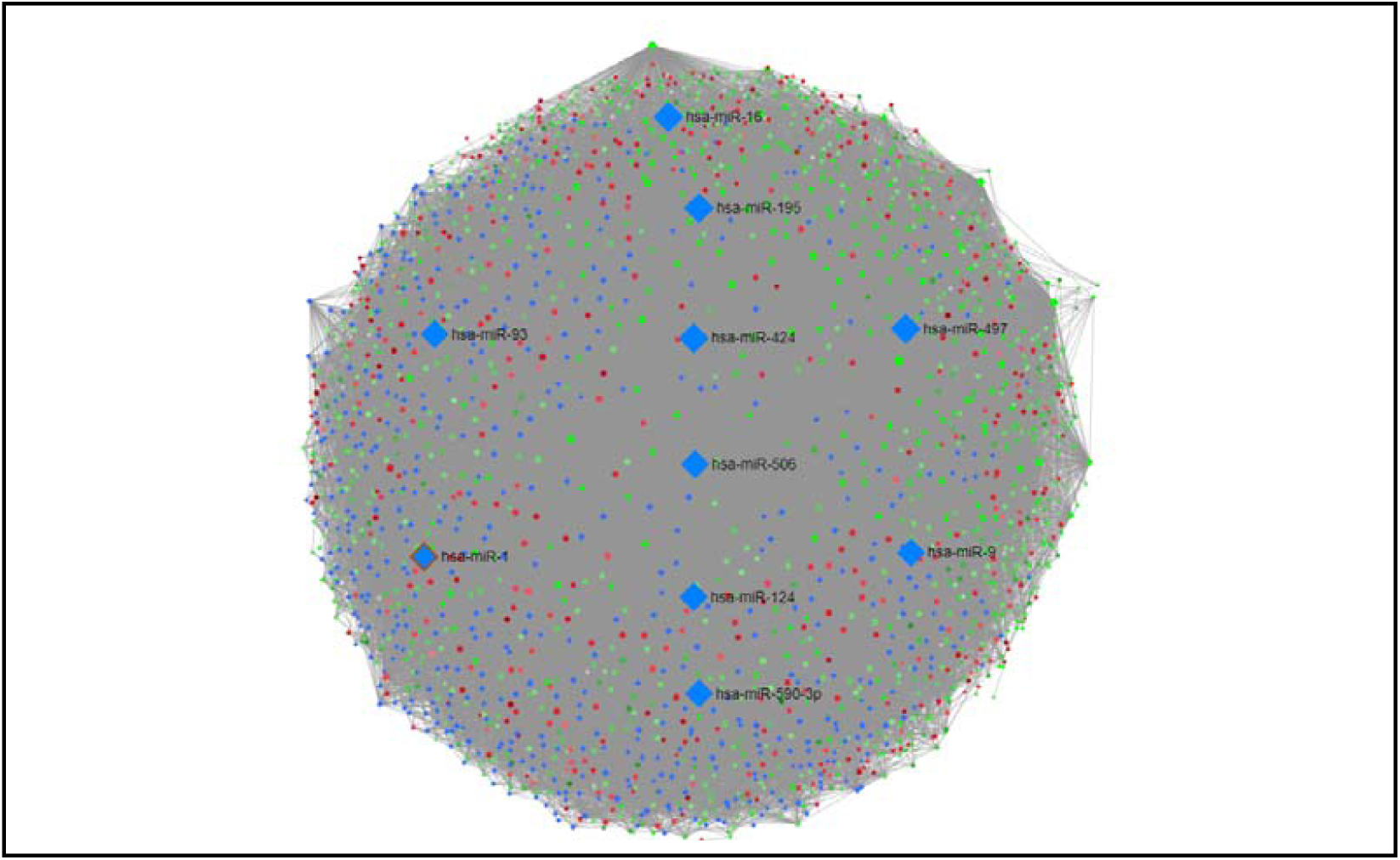
Interaction between miRNA and differentially expressed genes network of differentially expressed genes (DEGs). Nodes represent DEGs (Green: Upregulated genes; Red: Downregulated genes; Blue: miRNA). Edges represent interaction.

### 3.4. Identification of Therapeutic Candidate Molecules

Identified hub proteins and transcription factors were analyzed for their interaction with candidate molecules that could be used to reverse the PDAC condition. CREBBP, MAPK1, MAPK14, GRB2 and RPL23A among 10 hub proteins were reported to have significant interactions with candidate molecules (**Table 5**). Among the selected transcription factors, GATA2, PPARG and E2F1 showed interactions with multiple candidate molecules. Arsenic Trioxide, Dactinomycin, Bortezomib, Azactidine, Paclitaxel, Flourouracil and Carmustine were reported to be myelosuppressive agents. Anisomycin and Dactinomycin have nucleic acid synthesis inhibitory capability. Moreover, multiple candidate molecules were reported to be enzyme inhibitors.

**Table 5:**
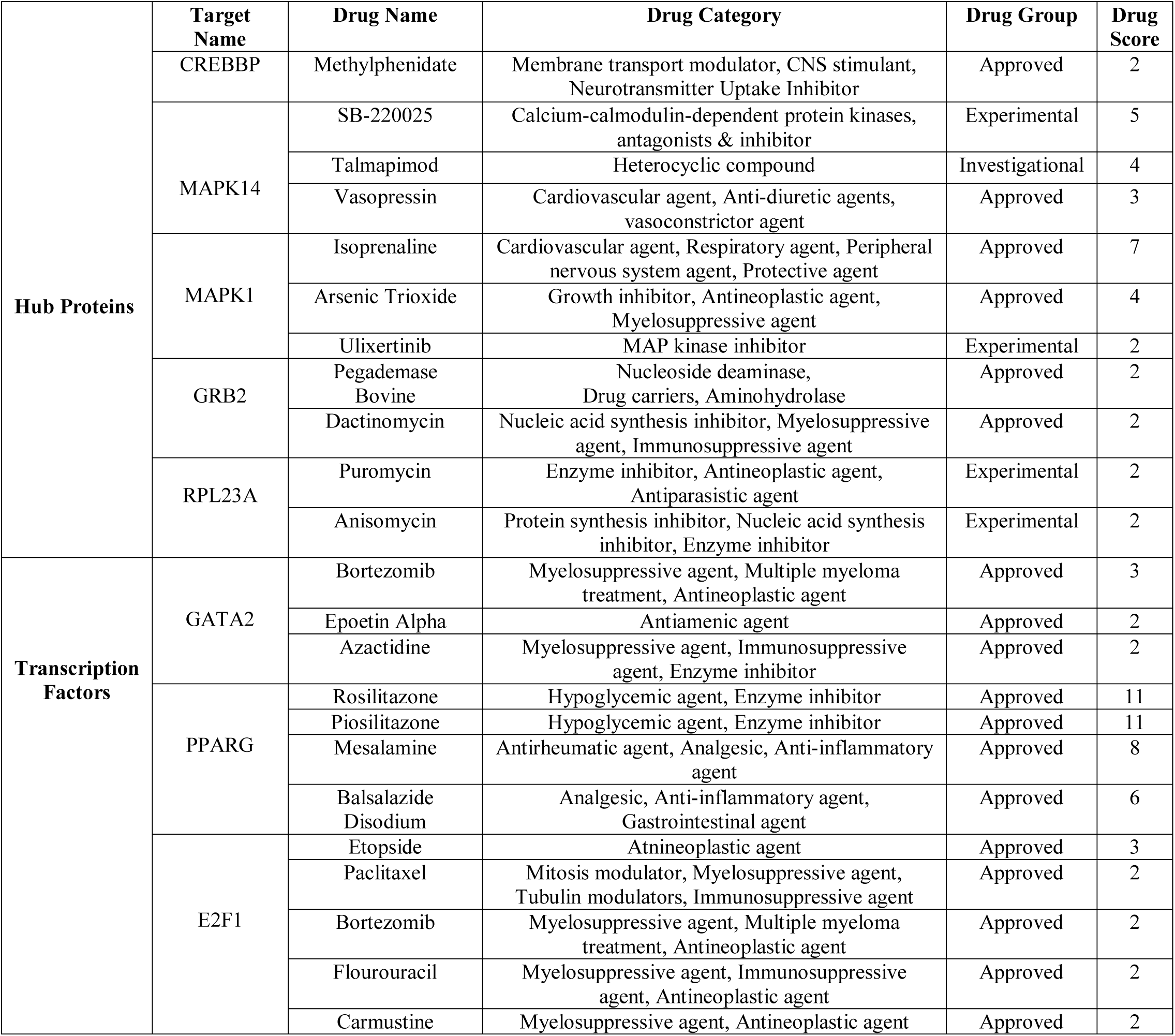
Summary of the selected candidate molecules on the basis of Drug-DEGs interaction.

|  | Target Name | Drug Name | Drug Category | Drug Group | Drug Score |
| --- | --- | --- | --- | --- | --- |
| Hub Proteins | CREBBP | Methylphenidate | Membrane transport modulator, CNS stimulant, Neurotransmitter Uptake Inhibitor | Approved | 2 |
|  | MAPK14 | SB-220025 | Calcium-calmodulin-dependent protein kinases, antagonists & inhibitor | Experimental | 5 |
|  |  | Talmapimod | Heterocyclic compound | Investigational | 4 |
|  |  | Vasopressin | Cardiovascular agent, Anti-diuretic agents, vasoconstrictor agent | Approved | 3 |
|  | MAPK1 | Isoprenaline | Cardiovascular agent, Respiratory agent, Peripheral nervous system agent, Protective agent | Approved | 7 |
|  |  | Arsenic Trioxide | Growth inhibitor, Antineoplastic agent, Myelosuppressive agent | Approved | 4 |
|  |  | Ulixertinib | MAP kinase inhibitor | Experimental | 2 |
|  | GRB2 | Pegademase Bovine | Nucleoside deaminase, Drug carriers, Aminohydrolase | Approved | 2 |
|  |  | Dactinomycin | Nucleic acid synthesis inhibitor, Myelosuppressive agent, Immunosuppressive agent | Approved | 2 |
|  | RPL23A | Puromycin | Enzyme inhibitor, Antineoplastic agent, Antiparasitic agent | Experimental | 2 |
|  |  | Anisomycin | Protein synthesis inhibitor, Nucleic acid synthesis inhibitor, Enzyme inhibitor | Experimental | 2 |
| Transcription Factors | GATA2 | Bortezomib | Myelosuppressive agent, Multiple myeloma treatment, Antineoplastic agent | Approved | 3 |
|  |  | Epoetin Alpha | Antiamenic agent | Approved | 2 |
|  |  | Azactidine | Myelosuppressive agent, Immunosuppressive agent, Enzyme inhibitor | Approved | 2 |
|  | PPARG | Rosilitazone | Hypoglycemic agent, Enzyme inhibitor | Approved | 11 |
|  |  | Piosilitazone | Hypoglycemic agent, Enzyme inhibitor | Approved | 11 |
|  |  | Mesalamine | Antirheumatic agent, Analgesic, Anti-inflammatory agent | Approved | 8 |
|  |  | Balsalazide Disodium | Analgesic, Anti-inflammatory agent, Gastrointestinal agent | Approved | 6 |
|  | E2F1 | Etoposide | Antineoplastic agent | Approved | 3 |
|  |  | Paclitaxel | Mitosis modulator, Myelosuppressive agent, Tubulin modulators, Immunosuppressive agent | Approved | 2 |
|  |  | Bortezomib | Myelosuppressive agent, Multiple myeloma treatment, Antineoplastic agent | Approved | 2 |
|  |  | Flourouracil | Myelosuppressive agent, Immunosuppressive agent, Antineoplastic agent | Approved | 2 |
|  |  | Carmustine | Myelosuppressive agent, Antineoplastic agent | Approved | 2 |

## 4. Discussion

After statistical analysis 1910 differentially expressed genes were identified with 681 upregulated and 1229 downregulated genes within the defined parameter which was then employed to analyze the functional enrichment.

Gene co-expression analysis and interaction network allows to identify functionally co-related genes, assume their tentative functions, identify regulatory biomolecules and understand disease-gene interactions (Van et al., 2017; Rhee et al., 2008). Upregulated and downregulated genes were found to be predominantly involved in translational process, apoptotic process, protein phosphorylation, immune responses, ATP binding, protein binding and signal transduction (**Table 1**). Protein phosphorylation is the most important post translational event in signal transduction which plays crucial role to mediate the cell signaling and its hyperactivity, malfunction or overexpression is mostly encountered in tumor and cancer development (Ardito et al., 2017). Altered apoptotic profile is a common clinical feature in pancreatic cancer (Bafna et al., 2009). PDAC involves an elevated level of both inflammatory and regulatory immune cells (Shibuya et al., 2014). Overexpression of ATP binding cassette protein has been observed in pancreatic cancer cells (Chen et al., 2012).

Protein-protein interaction (PPI) network provides useful insight into the functional organization of many proteins which promotes the understanding of complex molecular relationship to determine the phenotype of a cell (Stelzl and Wanker, 2006). Hub proteins are the most connected nodes in a PPI network that provides significant information about the function of the network (He and Zhang, 2006). Among, the selected 10 hub proteins, CREBBP is a transcriptional co-activator and its inhibition has been shown to suppress PDAC (Arensman et al., 2014) (**Table 2**). CREBBP has also been shown to play crucial role in lung cancer which was evident by its knockdown that resulted in the inhibition of lung cancer and induction of apoptosis (Tang et al., 2016). MAPK14 is a serine-threonine specific kinase that phosphorylates different proteins. MAPK14 dependent phosphorylation has been shown to mediate epithelial to mesenchymal transition in breast cancer cell (Hedrick and Safe, 2017). MAPK1 is another serine-threonine specific kinase and its overexpression has been reported in cervical cancer cells. MAPK1 has also been suggested to be constitutively active in absence of DUSP6 and leading to worsening prognosis of pancreatic cancer (Li et al., 2015; Furukawa, 2009). HSP90AB1 is a molecular chaperone and it promotes epithelial-mesenchymal transition in lung cancer (Wang et al., 2019). SMAD3 is a signal transducer and epithelial to mesenchymal transition in PDAC is mediated by upregulated SMAD3 (Yamazaki et al., 2014). STAT3, a signal transducer, whose overexpression has been shown to be correlated with VEGF (Vascular Endothelial Growth Factor) in pancreatic cancer. Constitutively activated STAT3 has been shown to be involved with elevated VEGF which then mediate angiogenesis and metastasis. Blockade of STAT3 showed suppression of VEGF expression (Wei et al., 2003). ACTB is a structural protein and it is essential for the maintenance of cytoskeletal structure and kinetics. 1.7 fold increase in expression of ACTB has been observed in pancreatic cancerous cells. However, ACTB overexpression has also been reported with other type of cancers i.e., lung cancer, colorectal cancer, liver cancer etc. (Rubie et al., 2005; Guo et al., 2013). MAGOH is a component of spliceosome and is required for splicing premature mRNA inside the cell. Differential expression of specific mRNA transcript has been correlated with MAGOH expression in breast cancer and this was evident by the knockdown of MAGOH along with other spliceosome factors which resulted in different pattern of mRNA production (Stricker et al., 2017). RPL23A is a component of ribosome and mediates the protein synthesis inside cell. Increased level of RPL23A expression has been observed in liver cancer (Molavi et al., 2019). UBC is responsible for ubiquitylation of many proteins inside cell. Downregulation of UBC has been shown to inhibit the proliferation of small cell lung cancer (Tang et al., 2015). Overexpression of UBC gene was observed in renal cancer cell (Kanayama et al., 1991).

Both transcription factors and microRNAs play crucial role in the expression of widespread genes inside human body (Martinez and Walhout, 2009). And the interactome network of these regulatory biomolecules provides crucial and valid information about the biological functions of these molecules and their involvement with disease (Vidal et al., 2011). Mostly connected 10 TFs and miRNAs were selected from TFs-DEGs and miRNA-DEGs interactome network (**Table 3**). Among the selected TFs, GATA2 has been reported to be overexpressed and accused as the metastasis-driving factor in prostate cancer (Chiang et al., 2014). High expression of both FOXC1 mRNA and protein has been observed in western blot and immunohistochemistry experiment with PDAC tissue (Wang et al., 2013). PPARG mutation has been observed in case of PDAC and some other form of cancers (Wang et al., 2015). Inhibition of PPARG by specific inhibitor has been shown to induce growth arrest of pancreatic carcinoma cells (Elnemr et al., 2000). E2F1 is essential for the S phase transition of PDAC cell (Schild et al., 2009). HINFP is responsible for regulating cell cycle and differential expression of this protein has been observed in different cancer cell lines (Holmes et al., 2005, van et al., 2009). Differential expression of USF-2 has been observed for bronchial dysplasia and non□adenocarcinoma lung cancer and this protein has been suggested as the early marker of lung cancer (Ocejo□Garcia et al., 2005). MEF2A has been reported to promote hepatocellular carcinoma (Pon and Marra, 2016). High expression of FOXL1 in PDAC tissue has been shown to promote the clinical outcome whereas lower expression is suggested to promote metastasis (Zhang et al., 2013). YY1 plays significant role in the signaling cascade that regulates the PDAC metastasis (Yuan et al., 2017). NFIC has been recently shown to play key role in breast cancer tumorigenesis (Lee et al., 2015).

Among the selected microRNAs, hsa-miR-93 is used as a biomarker for gastric cancer early detection and prognosis prediction (Larki et al., 2018) (**Table 4**). hsa-miR-16 is upregulated in PDAC and acts as a promising biomarker in cancer detection(Huang et al., 2019). hsa-miR-195 inhibits cell proliferation, migration, and invasion which potentially opens new avenues for the treatment of breast cancer(Singh et al., 2015). hsa-miR-424 is over expressed in PDAC patient and helps to characterize PDAC(Lee et al., 2007). hsa-miR-506 has downregulated expression profile in cervical cancer. Additionally, it works to promote apoptosis of cervical cancer cells (Li et al., 2011) hsa-miR-124 has been shown to be downregulated in nasopharyngeal carcinoma (NPC) and dramatically inhibited the cell proliferation, colony formation, migration and invasion in vitro, as well as tumor growth and metastasis in vivo (Peng et al., 2014). hsa-miR-590-3p can be used as a sensitive biomarker in coloreteral cancer. It is also tissue-specific and can regulate tumor suppressor genes and oncogenes in different tissues (Elfar et al., 2019). hsa-miR-1 is downregulated in the PDAC group compared with either in the sera samples or in tumor tissues. hsa-miR-1 was frequently decreased in clinical osteosarcoma (OS) tumor tissues and involved in the anticancer effect induced by specific chemical agent (Zhu and Wang, 2016). It also has a negative regulatory role in the proliferation of colon cancer by targeting baculoviral inhibitor of apoptosis protein(Xu et al., 2015). hsa-miR-497 is significantly downregulated in certain types of cancer, including breast, gastric, endometrial, colorectal and prostate cancer. It also inhibits both the migration and invasion of prostate cancer cells (Wu et al., 2016). hsa-miR-9 is up□regulated in non□small□cell lung cancer (NSCLC). It is also involved in transforming growth factor□beta 1 (TGF□β1)□induced NSCLC cell invasion and adhesion by targeting SOX7(Han et al., 2017).

DEGs were then analyzed to understand their potential interactions with small candidate molecule (**Table 5**). Among the selected candidate molecules, Arsenic Trioxide has been shown to induce apoptosis in pancreatic cancer cell (Li et al., 2003). Recently, Ulixertinib has been shown to have antitumor activity in PDAC in phase I clinical trial (Jiang et al., 2018). Anisomycin can decrease the proliferation of colorectal cancer cell (Ushijima et al., 2016). Bortezomib can induce apoptosis in PDAC cell (Nawrocki et al., 2005). Azactidine is capable of inducing apoptosis in ovarian cancer cell (Li et al., 2009).

Finally, publicly available microarray data of PDAC patient and control was used to identify potential biomarkers and drug targets using a network-based integrated approach. After continual computational assessment this study found several key proteome and regulatory signatures which may lead to the identification of potential biomarkers and drug targets. This study recommends CREBBP, MAPK14, MAPK1, SMAD3, UBC, MAGOH, HSP90AB1, RPL23A, ACTB and STAT3 as the best proteome signatures, GATA2, FOXC1, PPARG, E2F1, HINFP, USF2, MEF2A, FOXL1, YY1 and NFIC as the best transcriptional regulatory signatures, and hsa-miR-93, hsa-miR-16, hsa-miR-195, hsa-miR-424, hsa-miR-506, hsa-miR-124, hsa-miR-590-3p, hsa-miR-1, hsa-miR-497 and hsa-miR-9 as the best post-transcriptional regulatory signatures in PDAC patient. Moreover, identified signatures were also analyzed for their potential interactions with small candidate molecules. Anisomycin, Azactidine, Arsenic trioxide, Bortezomib, Ulixertinib and some other molecules were reported to have myelosuppressive agent that may reverse the PDAC condition.

## 5. Conclusion

PDAC is one of the most devastating form of cancers for which feasible treatment and early diagnosis techniques are merely available. In this study several genes have been identified which are predominantly involved in translational process, apoptotic process, protein phosphorylation, immune responses, ATP binding, protein binding and signal transduction in PDAC patient. Thereafter, several proteome and regulatory signatures were identified which were found to be involved in PDAC and some other form of cancers. Multiple therapeutic agents were also identified which may reverse PDAC condition. Other *in vitro* study also supported our findings and again we suggest further laboratory experiment to find the best potential biomarker and therapeutic agent for PDAC diagnosis and treatment. Hopefully, this study will raise research interest among researchers and contribute to the identification of feasible biomarker and drug target of PDAC in the upcoming days.

## Data Availability

All the data generated and analyzed are summarized in this manuscript.

## Acknowledgement

Authors are thankful to the members of Swift Integrity Computational Lab, Dhaka, Bangladesh.

## Data Availability Statement

All the data generated and analyzed are summarized in this manuscript.

